# Reemergence and global distribution of an invasive lineage of *Streptococcus pneumoniae* serotype 2

**DOI:** 10.64898/2026.03.13.26347380

**Authors:** Yogesh Hooda, Arif Mohammad Tanmoy, Krishna Banik Pushpita, Naito Kanon, Hafizur Rahman, Hakka Naziat, Harry C H Hung, Roly Malaker, Md Hasanuzzaman, Apurba Rajib Malaker, Deb Purna Keya, Sudipta Deb Nath, Belal Hossain, Shampa Saha, Mohammad Jamal Uddin, Keith P Klugman, Mathuram Santosham, Lesley McGee, Stephen D Bentley, Stephanie W Lo, Senjuti Saha, Samir K Saha

## Abstract

*Streptococcus pneumoniae* is a leading cause of childhood meningitis, sepsis and pneumonia despite widespread implementation of pneumococcal conjugate vaccines (PCVs). Serotype 2, once a major invasive serotype that nearly disappeared in the mid-20th century, is not included in current vaccine formulations. Recent reports from multiple countries suggest potential re-emergence of serotype 2. Here, we present 30 years of hospital-based surveillance from Bangladesh (1993-2022), where serotype 2 accounted for 7.8% of invasive pneumococcal disease cases. Infections occurred predominantly in very young infants (median age, 3 months) and were largely associated with meningitis (91.3%), with nearly 90% of isolates recovered from cerebrospinal fluid. Comparative analysis of otitis media and nasopharyngeal carriage isolates demonstrated high invasive propensity relative to other serotypes. Whole genome sequencing of 170 serotype 2 isolates from 21 countries revealed that all modern isolates belong to the globally disseminated lineage GPSC96, which is distinct from the prototypical laboratory strain D39 (GPSC622). Phylodynamic reconstruction dated the emergence of GPSC96 to the late 19th century, with continued global circulation and largely preserved antibiotic susceptibility. These findings highlight serotype 2 as a potential invasive pneumococcal threat in countries such as Bangladesh and supports consideration of its inclusion in the next-generation conjugate vaccines.

## Introduction

*Streptococcus pneumoniae* is a major cause of bacterial pneumonia, meningitis, and sepsis, leading to approximately 294,000 annual deaths among children aged 1–59 months globally in 2015 despite the widespread use of pneumococcal conjugate vaccines (PCVs)^1^. Two-thirds of these deaths occur in ten countries with high pneumococcal incidence, including Bangladesh, a sub-tropical country with a population of 170 million^2^. Since 2000, the widespread adoption of PCVs has had a major impact on the prevention of invasive pneumococcal diseases and severe pneumonia^3^. PCVs are projected to have prevented 1 million deaths among children worldwide by 2020, and are estimated to prevent an additional 6 million deaths by 2030^4^. The addition of serotypes that cause disease but are not contained in the currently available PCVs is likely to increase the impact of PCVs worldwide.

The World Health Organization (WHO) advocates for the inclusion of the PCVs in childhood vaccination schedules worldwide under national immunization programs (NIPs)^5^, noting large potential benefits in low- and middle-income countries that would adopt the vaccine^3^. Initially licensed in developed countries, PCVs have been found to significantly reduce invasive pneumococcal diseases (IPD) among children in several countries^6^. However, lack of comprehensive data on the vaccine’s impact in low- and middle-income countries (LMICs), coupled with the emergence of non-vaccine serotypes, can complicate assessments of PCV effectiveness in these regions. Consequently, the WHO has highlighted the need for ongoing monitoring of PCVs to fully understand their effectiveness in routine vaccination programs in countries with the highest disease burdens^4^. Currently, 174 of the 194 WHO member nations have introduced PCVs and 10 additional countries plan to introduce these vaccines.

Serotype 2 of *S. pneumoniae*, a major contributor of IPD cases in the early 20th century, saw a dramatic decline in cases starting in the 1940s and 1950s. However, by the early 1990s, records began to surface from some countries regarding increases in serotype 2. In Bangladesh, serotype 2 contributed to a 20% of all IPD cases with meningitis between 2002-2013^7,8^. In Israel, serotype 2 caused a significant outbreak beginning in 2015^9^. In Papua New Guinea (1989-2014), serotype 2 was a dominant cause of pneumococcal meningitis^10^. Barring these outbreaks, very few cases of serotype 2 have been reported between 2005-2019 in the WHO-commissioned multi-country Pneumococcal Serotype Replacement and Distribution Estimation (PSERENADE) surveillance study^11^. While not part of any approved PCV, serotype 2 has been included in upcoming pneumococcal conjugate formulations, which are currently undergoing clinical trials^12–15^.

This documented reemergence of serotype 2 in several countries necessitates a deeper understanding of its genomic and epidemiological characteristics. In this study, we describe demographic and clinical features of IPD cases caused by serotype 2 at four pediatric hospitals in Bangladesh from 1993 through 2022. Further, we describe the comparative genomic analysis of 170 serotype 2 isolates collected across 41 countries since 1989 and compare them with three ancestral serotype 2 strains from the early 20^th^ century. Collectively, our study highlights the continued importance of serotype 2 IPD and consideration for its inclusion in the next generation of pneumococcal vaccines.

## Methods

### Study design and isolate source

Pneumococcal isolates included in this study were obtained from multiple surveillance platforms in Bangladesh, including invasive pneumococcal disease (IPD) hospital surveillance, otitis media (OM) surveillance, and community-based nasopharyngeal (NP) carriage surveillance. Isolates were analyzed and compared by clinical syndrome and source.

### Invasive pneumococcal disease surveillance in hospitals

Invasive pneumococcal disease (IPD) surveillance has been ongoing at the Bangladesh Shishu Hospital and Institute (BSHI, formerly known as Dhaka Shishu Hospital, or DSH), the largest tertiary-care pediatric facility in Bangladesh, since 1993, conducted by the Child Health Research Foundation (CHRF). In 2004, three additional hospitals in Bangladesh were included in the IPD surveillance - Dr. M R Khan Shishu Hospital (MRKSH, formerly known as Shishu Shastho Foundation Hospital, or SSFH) in Dhaka, Chattogram Maa-O-Shishu (Mother and Children) Hospital, or CMOSH in Chittagong, and Kumudini Women’s Medical College and Hospital (KWMCH) in the rural town of Mirzapur. This surveillance has been part of the WHO-supported Invasive Bacterial Vaccine-Preventable Disease Surveillance Network^16^ and have been supported by different projects at different times. Children with suspected IPD infections based on clinical examination by study physicians were enrolled into the surveillance upon written consent from the legal guardian/parents. Age, demographic, hospital outcome data was collected and stored from all cases.

#### Case definition of IPD

Specimens (blood and/or CSF) for identification of IPD were collected from children at the discretion of the treating physician. Those who met the case definition of pneumonia, severe pneumonia, meningitis, and/or sepsis according to the WHO were selected for this study^8^. The following case definitions were used by the study physicians of all four hospitals in the surveillance network to identify children with suspected pneumococcal disease and blood cultures were obtained from them for confirmation of IPD:

##### Pneumonia

Pneumonia was defined as history of cough or breathing difficulty with fast breathing (respiration rate ≥ 50/min for children 2- <12 months of age, ≥ 40/min for children ≥ 12 months of age).

##### Severe pneumonia

A case of severe pneumonia was defined as history of cough or breathing difficulty and presence of any of the following danger signs (chest indrawing, inability to drink/breast feed, substantial vomiting, convulsions, lethargy, or fast breathing (respiratory rate ≥ 60 breaths/min in children < 2 months of age), stridor in a calm child, altered consciousness).

##### Meningitis

A case of meningitis was defined as sudden onset of fever (>100.4°F) and presence of at least one of the following: neck stiffness, altered consciousness, bulging fontanel if < 12 months of age, lethargy, convulsions, toxic appearance, petechial/purpural rash, poor sucking, or irritability.

##### Sepsis

A case of sepsis was defined as presence of any of the following danger signs (inability to drink/breast feed, convulsions, lethargy, stridor in a calm child, persistent vomiting, or severe malnutrition) excluding pneumonia and meningitis features.

### Otitis media (OM) surveillance amongst hospitalized children

CHRF conducted OM surveillance at BSHI hospital in Dhaka, Bangladesh during 2014-2019. The study screened children under 18 years who came to the hospital’s Ear, Nose, and Throat (ENT) outpatient department. The study enrolled children with suspected OM, identified by study physicians following clinical and otoscopic criteria recommended by the WHO. More details about the surveillance have been described in earlier work^17,18^.

### Nasopharyngeal carriage surveillance in the community

NP carriage surveillance was conducted among healthy children in eight unions of Mirzapur, Tangail, approximately 60 km north of Dhaka city, Bangladesh^19^. The objective was to determine the prevalence of pneumococcal carriage among healthy children in the community. Between 2014 and 2016, a total of 1,576 nasopharyngeal swabs were collected from children aged 4-36 months by trained healthcare workers following informed consent from a parent or legal guardian. Children were randomly selected from the community using a computer-generated random list created annually for each age group from the Mirzapur health and demographic surveillance system (MHDSS) database^20^, based on unique identification numbers.

### Microbiological methods and serotyping

All blood/CSF specimens from the IPD study were cultured on McConkey, blood- and chocolate agar plates. Since 2016, blood cultures were performed using BACTEC^TM^ automated blood culture system (Becton, Dickinson and Company, USA). Beep-positive and/or chocolate-colored blood bottles were subsequently screened by culturing on blood- and chocolate agar plates.

All ear discharge specimens (OM study) and nasopharyngeal swab specimens (NP study) were cultured on selective media including blood agar, chocolate agar, and MacConkey agar.

The suspected *S. pneumoniae* colonies from all plates were confirmed based on their susceptibility to optochin (5 mg). Isolates with an inhibition zone (diameter) of > 14 mm were confirmed as *S. pneumoniae*. Isolates with a zone of 9–13 mm were subjected to a bile solubility test for further confirmation^21^.

Pneumococcal isolates were serotyped using capsular swelling method (Quellung reaction). Since 2004, all CSF specimens that showed no growth on plates underwent testing for pneumococcus using an immunochromatographic method - BinaxNOW *S. pneumoniae* antigen card assay (Abbott Laboratories, Chicago, IL, USA)^22^. All BinaxNow-positive specimens were tested for the *lytA* gene of pneumococcus and serotyped using previously published seven multiplex (triplex) and three in-house multiplex (triplex) RT-PCR assays covering a total of fifty serotypes (1, 2, 3, 4, 5, 6A/6B/6C/6D, 6C/6D, 7A/7F, 7B/C, 8, 9 V/9A, 10A/F, 11A/D, 12A/12B/12F/44/46, 13, 14, 15A/B/C/F, 15A/15F, 16F, 18A/18B/18C/18F, 19A, 19F, 20, 22A/22F, 23A, 23F, 33A/33F/37, 34, 35B, 45 and 48)^21,23^.

Presence of antibiotic in the CSF was tested using a method described previously^23^. Briefly, a blank disc (product CT998; Oxoid) was placed on a lawn of the pan-susceptible organism *Micrococcus luteus* (American Type Culture Collection 9341). A 10-μL CSF sample with ≥10 WBC/μl was pipetted directly on the blank disc before incubating the plate overnight at 37°C. Any plate with a zone of inhibition around the disc was considered positive for the presence of an antibiotic.

### DNA extraction & whole-genome sequencing

For all serotype 2 pneumococcal isolates, DNA was extracted from multiple single colonies using an optimized protocol of the Qiagen DNA Mini Kit (Qiagen, Hilden, Germany). The protocol includes a pre-incubation phase (60 minutes at 37°C, followed by 30 minutes at 56°C) with enzyme mixes containing lytic enzyme and proteinase K. WGS was conducted under the Global Pneumococcal Sequencing Project (GPS), and GPS2. In the initial phase of the GPS project, sequencing was performed on the Illumina Hiseq or Novaseq platform at the Wellcome Sanger Institute, Cambridge, UK. More recently, under the GPS2 project, sequencing was carried out on the Illumina NextSeq2000 platform at the CHRF Genomic Center in Dhaka, Bangladesh. The sequencing read length for all platforms was 150-bp paired-end. In total, 67 isolates were sequenced with GPS (timeline: 2003-2017) and one with the GPS2 project (timeline: 2022).

### Serotype confirmation and detecting Global Pneumococcal Sequence Clusters (GPSCs), Sequence Types (STs), and resistance genes

All raw fastq files were transferred to the CHRF server and underwent downstream analysis for quality assessment and trimming, utilizing FastQC v0.11.5 and fastp v0.23.2^25^. De novo genome assembly and annotation were performed using Unicycler v0.4.9^26^ and Prokka v1.14.6^27^, respectively. The serotype confirmation, antimicrobial resistance (AMR) genes, GPSCs and STs were identified using the GPS Pipeline version-rc11 (https://github.com/GlobalPneumoSeq/gps-pipeline) for all genome data^28^.

For reference-based genome mapping, Bowtie2 was used to map all genome data, with the complete chromosome of a serotype 2 strain D39 (accession: CP000410.2) serving as the reference^29^. SAMtools and BCFtools were employed for variant calling, with candidate Single Nucleotide Polymorphisms (SNPs) filtered for a minimum base quality of 20, a minimum high-quality read count of 5, and >75% read support (>75% of the reads supporting the SNP over the reference)^30^. SNPs were also filtered to eliminate homogeneity and biases, including strand bias, mapping bias, and end bias (p-value < 0.001). SNPs located in phage regions (as detected by Phaster), repeat sequences, and recombination regions (as detected by Gubbins) were excluded from the genome^31^. The final variant vcf file, containing the filtered SNPs, was then converted to a SNP alignment which was used by RAxML to construct a phylogenetic tree^32^. Tree visualizations of subtypes, sample sources, GPSCs, STs, and years were achieved using iTOL v6^33^.

### Hybrid assembly and reconstruction of the cps loci

To reconstruct the complete sequence of the capsular polysaccharide synthesis (*cps*) loci of a serotype 2 GPSC96 isolate (SPN_0164; collected in 2022), we performed a hybrid assembly of Illumina and Oxford Nanopore Technology (ONT) reads using Unicycler v0.5.1. The resulting contigs were then scaffolded against the serotype 2 reference chromosome (strain D39; accession: CP000410.2) with RagTag v2.1.0^34^, inserting 100-bp of Ns between contigs to generate a pseudo-complete chromosome. Next, this sequence was examined to identify the *dexB* and *aliA* genes, which flank the *cps* loci, based on the serotype 2 reference genome (NCBI accession: NC_008533.2). The sequences of *dexB*, *aliA*, and the intervening region were extracted and aligned to the reference genome using Blastn to define the complete *cps* loci. Annotation was performed using PGAP (version 2024-07-18.build7555; https://github.com/ncbi/pgap)^35^, with gene names assigned according to the serotype 2 *cps* loci reference from Bentley et al 2006^36^. Using the same method, the *cps* loci from the GPSC622 reference genome (NCBI accession: NC_008533.2) was also extracted and annotated.

### Global collection of sequenced serotype 2 genomes

Global *S. pneumoniae* serotype 2 genomes were collected from the Monocle (https://data-viewer.monocle.sanger.ac.uk/project/gps, date: January 24, 2025), screened using the same GPS pipeline, filtering through the “InSilicoSerotype = 2”. In total, 102 genomes were identified, from which country and year of isolation data were available for 99 and 92 genomes respectively (Supplementary data 1). No additional serotype 2 genomes were obtained from PubMLST using search “Serotype=2”. Apart from the global collection, 68 Bangladesh genomes sequenced in this study were added to the database. In total, 170 genomes were obtained from 21 countries, of which 3 genomes had no country information.

### Phylodynamic analysis using BEAST

Bayesian phylodynamic analyses were conducted to investigate the global collection of serotype 2 isolates using BEAST v1.10.4^37^. The substitution model was determined to TVMe under the Bayesian information criterion by ModelFinder^38^. The exponential unrelated relaxed clock as clock type and Bayesian skyline coalescent model as tree prior were used. The analysis considered the year of isolation as tip dates and continued for 500 million steps with sampling every 25,000 iterations. An asymmetric discrete trait phylodynamic modeling approach was used to incorporate countries of isolation. The BEAST analysis was run three times. The resulting log files and model parameters were analyzed on Tracer v1.7.1^39^. TreeAnnotator v1.10 was used to generate the maximum–clade-credibility (MCC) tree. The tree was visualized and annotated using iTOL v6^40^. The model with the highest posterior values (joint effective sample size (ESS) of 3631 was used for annotation and further analysis.

### Data Analysis

Data analysis and visualization were conducted using Python (version 3.10.12) within the JupyterLab environment (version 4.0.7). The python libraries, *NumPy* and *Pandas* supported the analytical processes, while *GeoPandas* was used for mapping global data. Visualization was done using the *Seaborn* and *Matplotlib* libraries. In addition, visualization of gene comparison within the capsular (*cps*) loci between GPSC96 and GPSC622 was performed using the *gggenes* package v0.5.1 in R.

## Results

### Yearly distribution of IPD cases attributable to serotype 2 in Bangladesh

Between 1993 and 2022, *S. pneumoniae* was identified in 2,228 children, with 7.8% (173/2228) cases classified as serotype 2. Until 2004, diagnosis was based on CSF or blood cultures. Since then, an immunochromatic test (ICT) was added in 2004 for detecting *S. pneumoniae* in CSF, followed by PCR for serotyping^23^. Since 2014, 92% (36/39) of serotype 2 cases have been detected using ICT, while 8% (3/39) were identified through blood cultures (Table 1). Among the 69 cases, 58 had prior antibiotic exposure, with 49 (84.4%) being culture-negative and only 9 (15.6%) culture-positive. In contrast, of the 11 cases without any prior antibiotic exposure, 6 (54%) were culture-positive and 5 (46%) were culture-negative. These findings suggest that serotype 2 is increasingly being detected through culture-negative methods, primarily ICT in CSF, likely due to increases in prior antibiotic exposure before a patient reaches the tertiary level hospital.

**Table 1:**
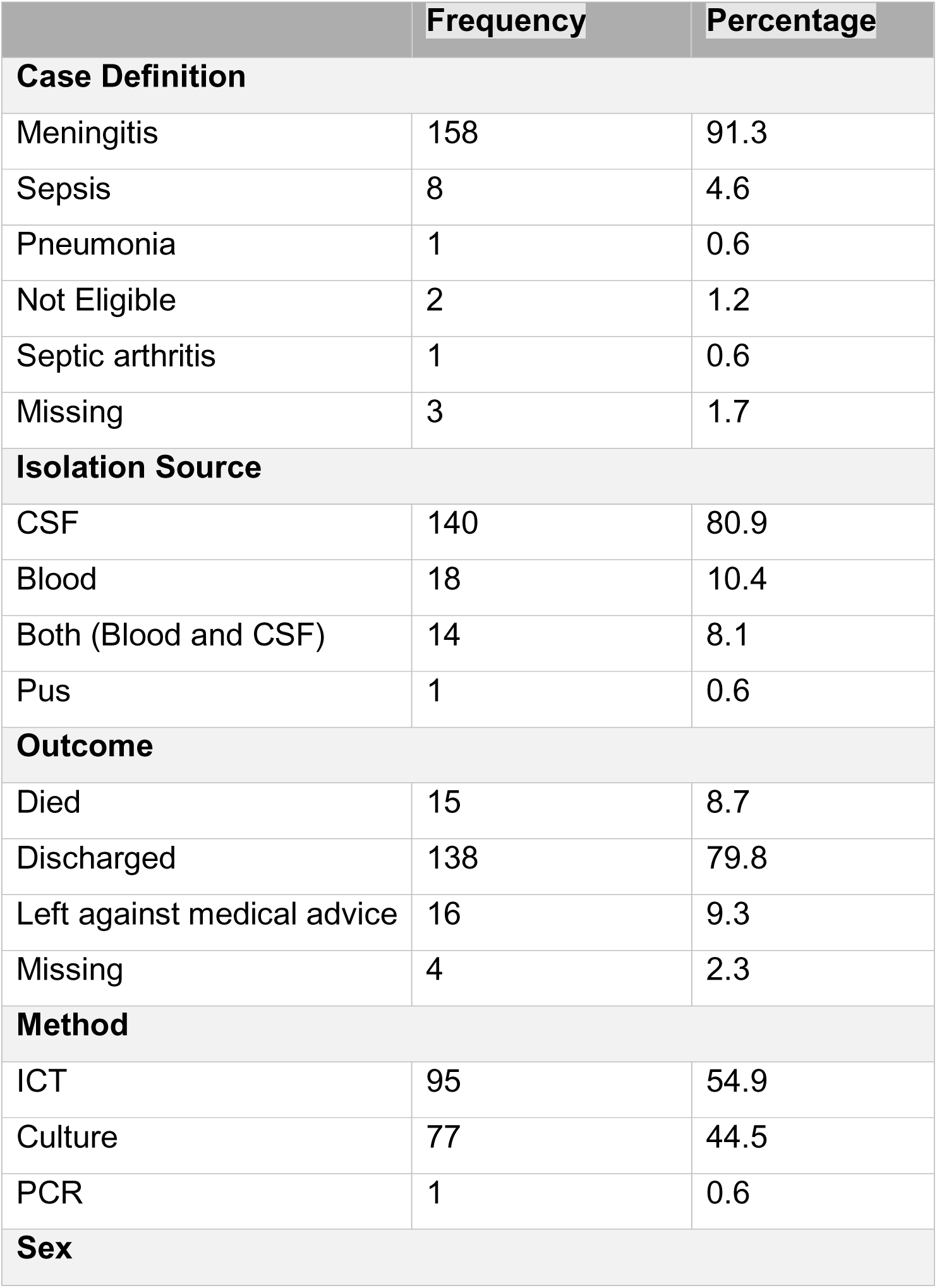
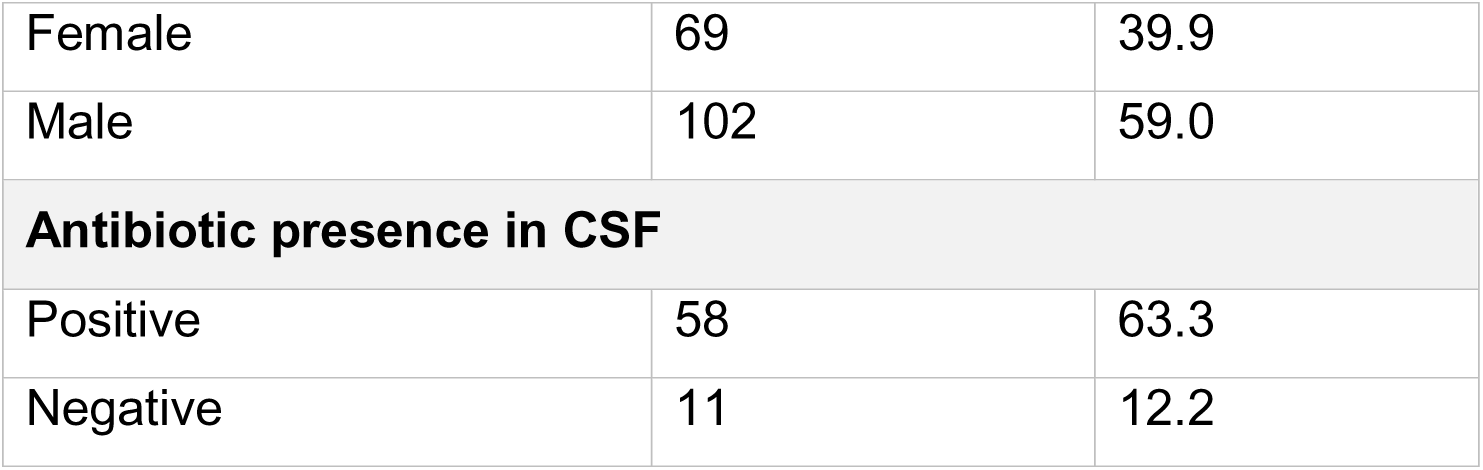
Demographic and clinical characteristics of invasive pneumococcal infections in Bangladesh associated with serotype 2.

|  | Frequency | Percentage |
| --- | --- | --- |
| **Case Definition** |  |  |
| Meningitis | 158 | 91.3 |
| Sepsis | 8 | 4.6 |
| Pneumonia | 1 | 0.6 |
| Not Eligible | 2 | 1.2 |
| Septic arthritis | 1 | 0.6 |
| Missing | 3 | 1.7 |
| **Isolation Source** |  |  |
| CSF | 140 | 80.9 |
| Blood | 18 | 10.4 |
| Both (Blood and CSF) | 14 | 8.1 |
| Pus | 1 | 0.6 |
| **Outcome** |  |  |
| Died | 15 | 8.7 |
| Discharged | 138 | 79.8 |
| Left against medical advice | 16 | 9.3 |
| Missing | 4 | 2.3 |
| **Method** |  |  |
| ICT | 95 | 54.9 |
| Culture | 77 | 44.5 |
| PCR | 1 | 0.6 |
| **Sex** |  |  |
| Female | 69 | 39.9 |
| Male | 102 | 59.0 |
| **Antibiotic presence in CSF** |  |  |
| Positive | 58 | 63.3 |
| Negative | 11 | 12.2 |

Our analysis highlights a fluctuating trend in the number of cases, with an initial period of relatively low incidence from 1996 to 2003 (Fig 1). The number of serotype 2 cases subsequently increased between 2004 and 2011. After 2011, a noticeable decline was observed, leading to a stabilization at a lower yet consistent annual case count. Since serotype 2 was not included in PCV10, the vaccine used in Bangladesh, there were no major changes observed between the pre-PCV10 era (1993-2014), where serotype 2 accounted for 8.35% (136/1628) of non-PCV10 serotypes, and the post-PCV10 period (2015-2022), where it accounted for 6% (36/600) of all non-PCV10 serotypes.

**Figure 1:**
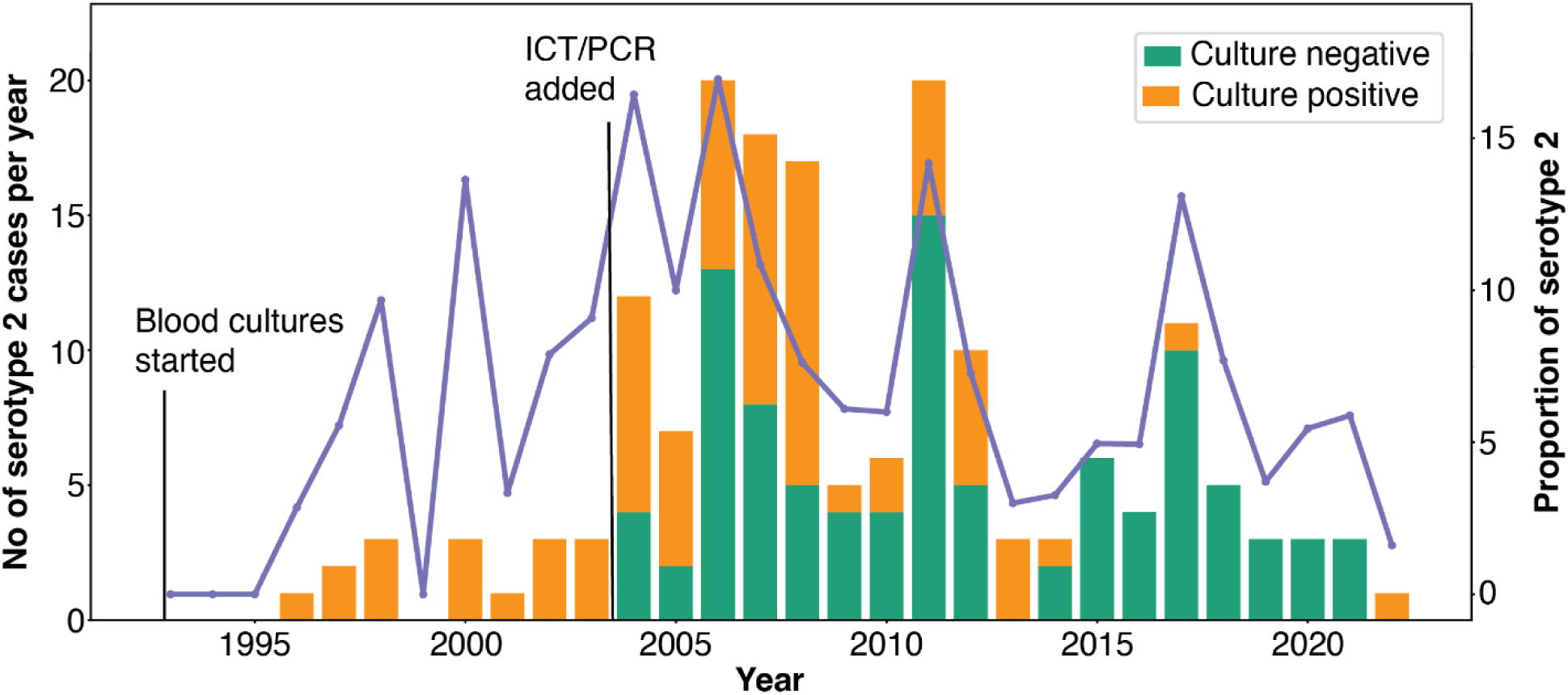
Temporal trends in *S. pneumoniae* serotype 2 isolates from invasive pneumococcal disease cases in Bangladesh, 1993-2023. Annual counts of serotype 2 isolates (left y-axis) colored based on detection method (orange: detection through blood culture, green: detection through culture-negative methods: immunochromatic test (ICT) or PCR). Their proportion relative to all pneumococcal isolates (purple line, right y-axis) are shown. Serotype 2 is the second most common IPD serotype in Bangladesh (173/2228, 7.8%).

We also investigated the prevalence of serotype 2 among *S. pneumoniae* isolates from otitis media (OM) cases at the Bangladesh Shishu Hospital from 2014 to 2019, as well as nasopharyngeal swabs from healthy children in the community from 2014 to 2018. In total, serotype 2 was identified in 0.4% (3/745) of S. *pneumoniae* isolates from OM cases, while it was not detected at all (0/1421) from nasopharyngeal carriage samples. To assess the invasiveness potential of serotype 2, we compared the proportions of various pneumococcal serotypes in cases of IPD versus OM cases (Fig. 2a). Serotype 2 was found to be responsible for a higher proportion of invasive infections, similar to other serotypes known for their high invasive potential that are covered by existing PCVs. A comparison of the proportion of all different pneumococcal serotypes in IPD and nasopharyngeal carriage cases also yielded similar results (Fig 2b). Collectively, these findings indicate the high invasive potential of serotype 2 strains in Bangladesh.

**Figure 2:**
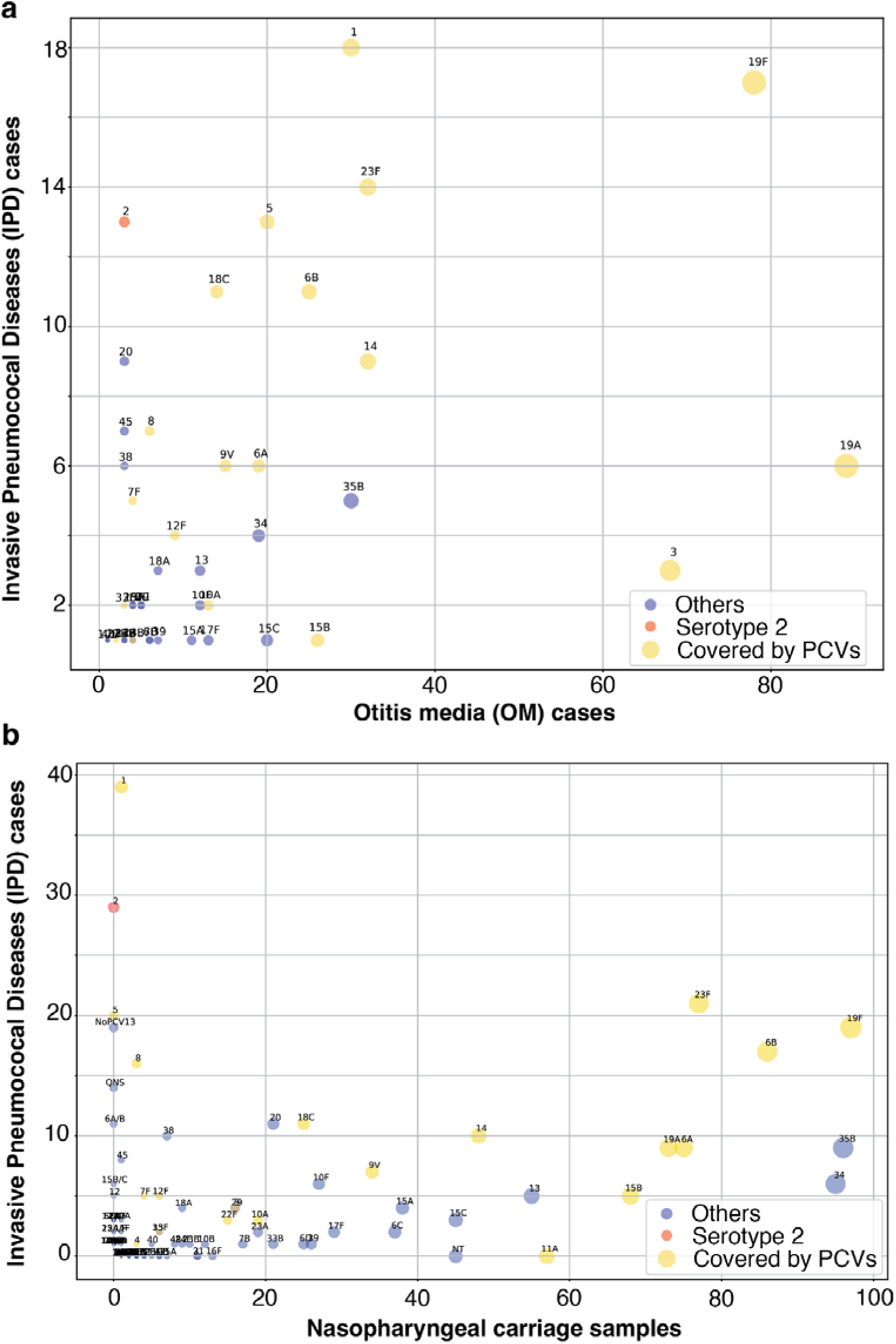
The relative proportion of different serotypes amongst invasive pneumococcal disease cases (y-axis) vs a) otitis media cases (x-axis), or b) nasopharyngeal carriage (x-axis). Cases between 2014-2019 are shown. The size of points indicates the number of cases in IPD. Serotype 2 has been colored in red; all serotypes present in WHO-approved pneumococcal conjugate vaccines (PCVs – PCV7, 10, 13, 15 and 20) are colored in yellow. All non-PCV and non-2 serotypes are shown in blue.

### Demographic and clinical characteristics of cases with serotype 2 infections

The median age for cases of IPD caused by serotype 2 was 3 months, compared to 8 months for IPD cases attributed to other *S. pneumoniae* serotypes (Fig 3). The mortality rates for serotype 2 cases (8.67%; 16/173) and non-serotype 2 cases (8.16%; 182/2228) were similar(Table 1).

**Figure 3:**
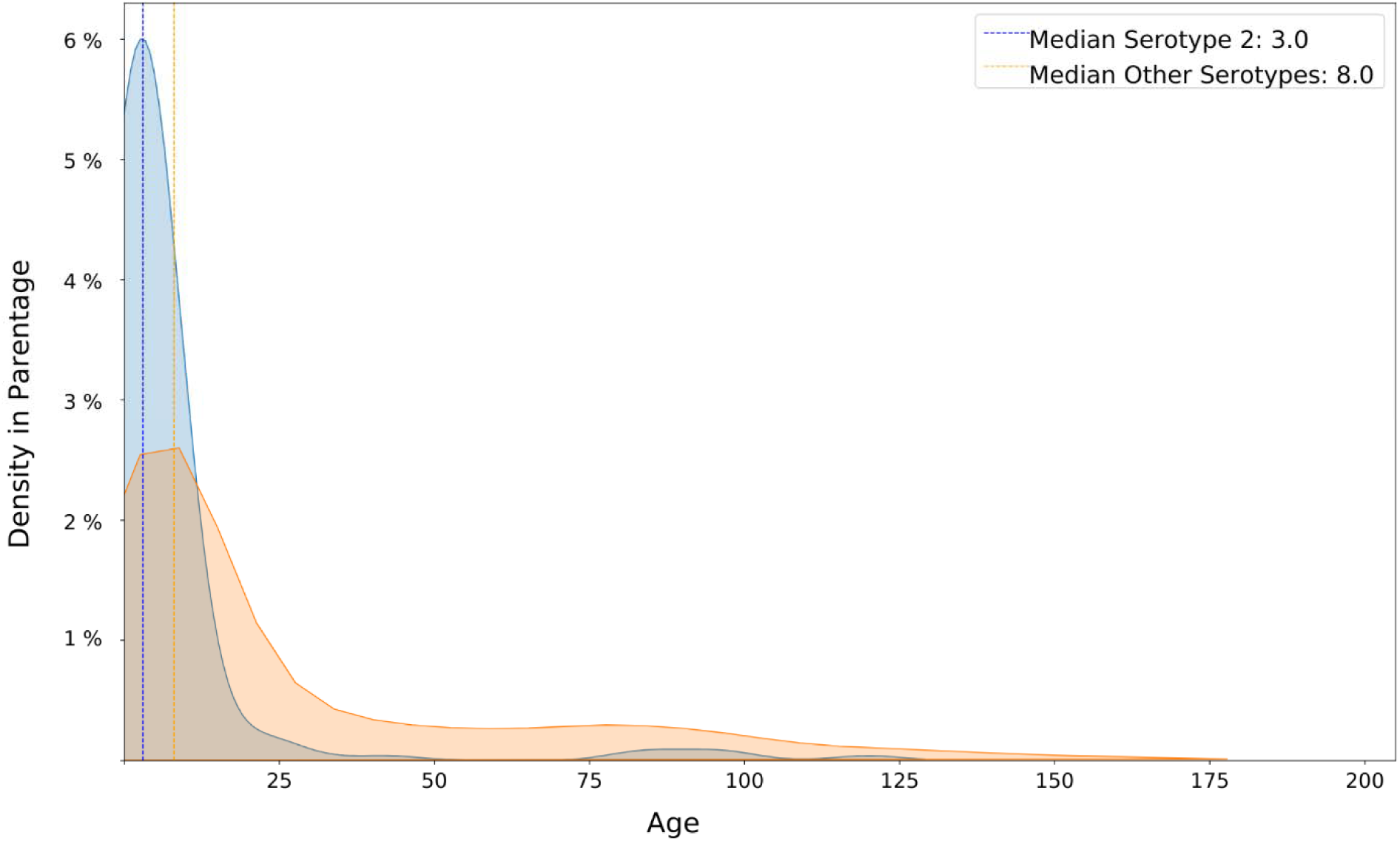
Age distribution of invasive pneumococcal disease (IPD) caused by serotype 2 (blue) and other serotypes (orange) in Bangladesh. The median age for serotype 2 IPD was 3 months (95% CI: 3.97 to 8.94), while the median age for IPD cases caused by other serotypes is 8 months (95% CI: 3.97 to 8.94). Density curves represent kernel density estimates of age distribution.

The analysis of the sampling source revealed that serotype 2 infections demonstrated a higher propensity to be neuroinvasive compared to non-serotype 2 infections. Specifically, 89.02% (154/173) of serotype 2 infections were detected from CSF, compared to 77.15% (1719/2228) for all other serotypes (Table 1). Meningitis was identified as the primary disease associated with serotype 2, occurring in 172 out of 173 infections (99.42%), compared to 2146 out of 2228 (96.32%) for non-serotype 2 infections (Table 1). Furthermore, among culture-positive cases (from both CSF and blood), the proportion of serotype 2 infections resulting in meningitis was higher than that of non-serotype 2 infections (Table 1).

### Global phylogenetic analysis of serotype 2 isolates

To assess the genetic diversity of serotype 2, we performed whole-genome sequencing on isolates identified from positive blood cultures in Bangladesh. Out of 77 culture-positive isolates previously biobanked, we successfully revived and sequenced 68 isolates. These sequenced isolates were obtained between 2003 and 2022, spanning 20 of the 30 years of the surveillance period. All strains from Bangladesh belonged to GPSC-96, with ST-74 being the most frequent (46/68, 67.6%) (Fig 4).

**Fig. 4:**
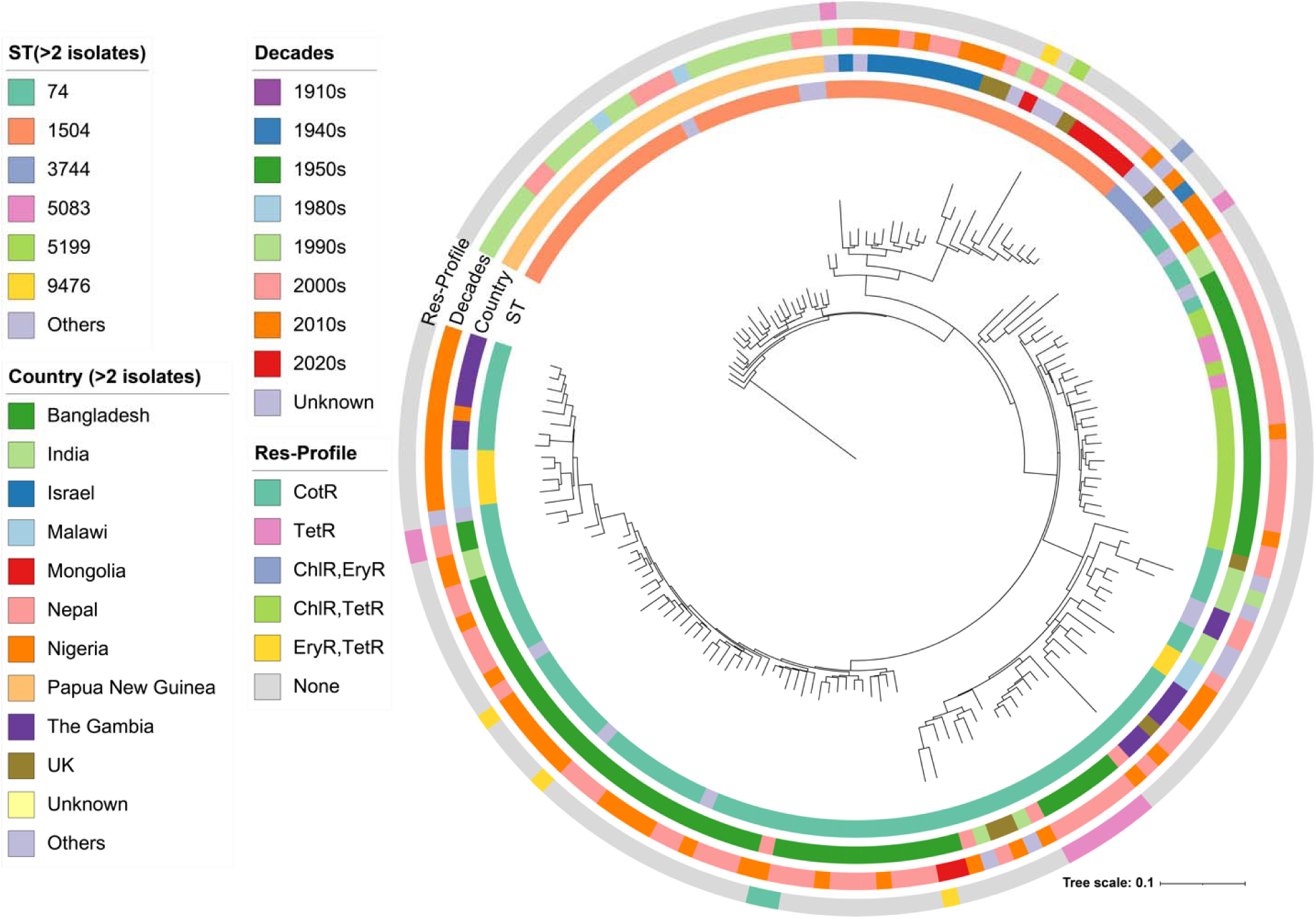
A maximum-likelihood-based phylogeographic tree of global serotype 2 isolates. The country of isolation, GPSC, ST, decade of isolation and resistance profile (Res-Profile) is shown in the rings around the phylogenetic tree. The Res-Profile circle is showing resistance (by the presence of genes/mutations, following the results from GPS pipeline) to Cotrimoxazole (CotR), Tetracycline (TetR), Chloramphenicol (ChlR), and Erythromycin (EryR).

To contextualize the serotype 2 genomes from Bangladesh, we searched the Monocle^42^ and PubMLST^43^ datasets to identify 102 additional serotype 2 isolates collected between 1916 and 2017, excluding those from Bangladesh. Due to the scarcity of available isolates and WGS data, very few isolates were obtained prior to 1989 (n=3; 1916, 1943 and 1956). Since then, 99 serotype 2 genomes were identified in 18 countries including Papua New Guinea (n=27), Gambia (n= 14), India (n=11), Israel (n=9), UK (n=8), Malawi (n=6), and 11 other countries or unknown origin (n=24). All of these genomes were sequenced as part of the Global Pneumococcal Sequencing (GPS) project^44^. In contrast to Bangladesh, the source of isolation of global serotype 2 isolates (n=102) was more diverse including CSF (n=41), blood (n=27), nasopharynx (n=6), blood/CSF (n=4), sputum (n=1), ear discharge (n=1), and unknown sources (n=22).

Three distinct GPSCs were identified among the isolates; 96 (n=166), 49 (n=2), and 622 (n=2), with GPSC96 being the predominant cluster (Sup Fig. 1). Notably, all strains analyzed from post-1989 belong to GPSC96. The oldest sample within GPSC96, dating back to 1943, originated from Denmark and with an ST-3744 (Fig 4). These findings suggest that, among the three circulating lineages from the early 20^th^ century, only GPSC96 continues to circulate today.

Among the 170 serotype 2 isolates assessed for antimicrobial susceptibility to 19 common antibiotics predicted using the GPS pipeline, the majority (88%; 150/170) were susceptible to all tested antibiotics (Fig 4). Among the treatment options, only 6 isolates (3%) were erythromycin-resistant (EryR) while no penicillin resistance (PenR) was observed. Tetracycline resistance (TetR) was found in 9% (15/170; conferred by the *tetM* gene) of the isolates. Instances of combined resistance profiles were rare, with “EryR/TetR” observed in 2% (4/170) isolates, chloramphenicol + erythromycin resistance (ChlR/EryR) in 0.6% (1/170), and chloramphenicol + tetracycline resistance (ChlR/TetR) in 0.6% (1/170). For the dominant lineage GPSC96, 89% (148/166) showed no resistance, closely reflecting the overall population (89%; 150/170). Two distinct genetic determinants for EryR were identified; primarily the macrolide efflux genes, *mefA-msrD* (n = 5; all from GPSC96), and *mefE* (n = 1; from GPSC622).

To investigate the differences between the currently circulating clinical strain (belonging to GPSC96) and the laboratory reference strain (belonging to GPSC622), we compared the *cps* loci using one representative sequence from each GPSCs. We generated long-read genome data by Nanopore sequencing for the most recent Bangladeshi GPSC96 isolate in the database (Sample_ID: SPN_0164, from 2022). A hybrid assembly with its Illumina data resulted in a near-complete genome (2,052,236 bp; four contigs). Using this assembled genome, a pseudo-complete chromosome (2,052,536 bp, incl. 300 bp of Ns in between the contigs) was generated using RagTag (see Methods). This analysis revealed a *cps* locus of 23,631 bp in GPSC96, compared to 23,907 bp in the GPSC622 reference genome (NCBI accession: NC_008533.2) (Supplementary Fig. 2). While the overall *cps* gene content and sequences were conserved in both GPSCs, differences were noted in the transposon regions adjacent to *dexB* and *aliA* genes. Specifically, in GPSC96, a *tnp630* transposon located next to *dexB* was absent, and three *IS66* elements adjacent to *aliA* were replaced by pseudogenes, *glf_2* and *glf_3* (Supplementary Fig. 2).

### Global distribution of serotype 2 isolates

Different serotype 2 ST’s belonging to GPSC96 were mapped onto a world map to illustrate the global distribution of serotype 2 genomes (Fig 5). Notably, three distinct clades were identified across a variety of countries. The clade belonging to ST74, 5199, 9476, and 5083 was predominantly found in Bangladesh, India, Nepal, and The Gambia. The clade with ST1504 was observed in Papua New Guinea, Israel, and Mongolia. Additionally, the 3 genomes belonging to ST3744 were in Argentina, Malawi, and Denmark. The distribution of these STs across different countries suggest a lack of a common source for the increase in serotype 2 cases observed post-1989.

**Figure 5:**
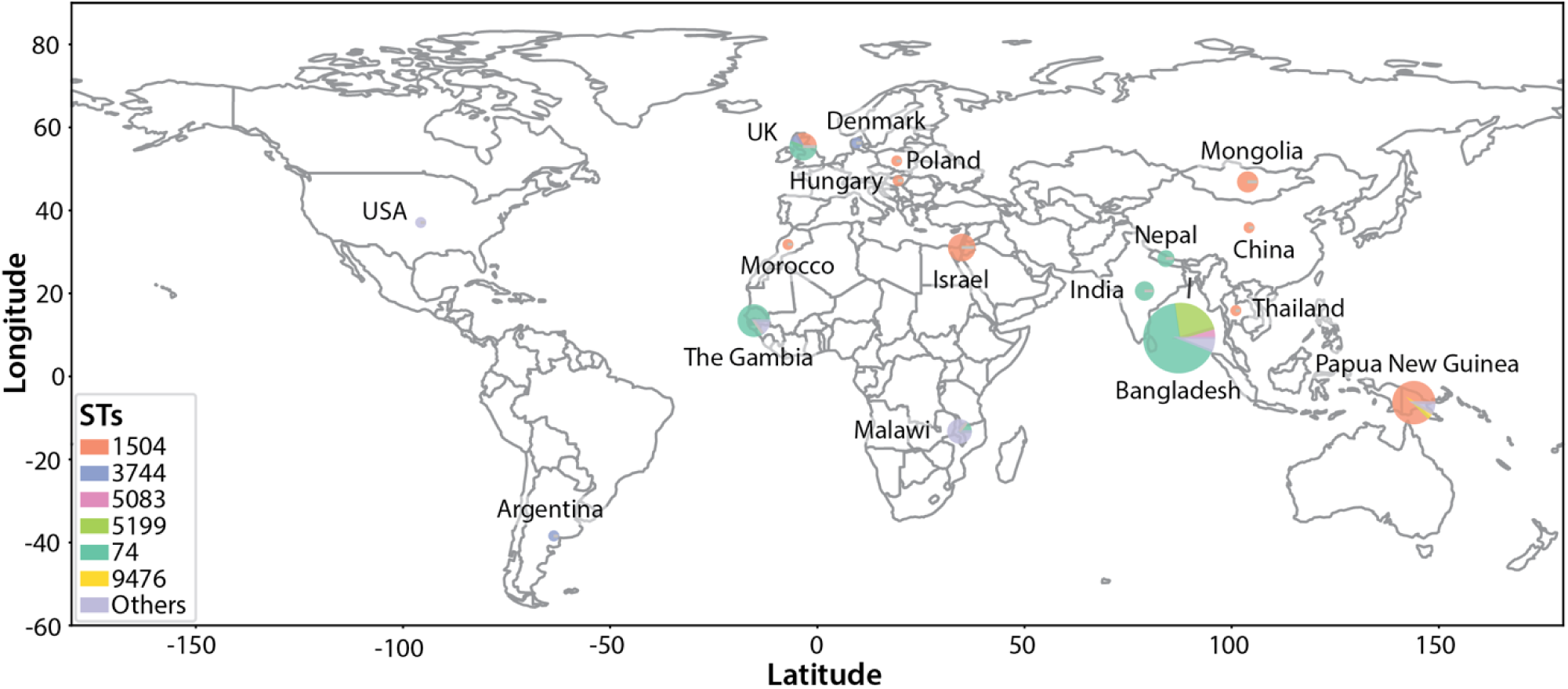
Geographical distribution of serotype 2 genomes from 1916-2022. Circles correspond to the number of genomes. Each circle is colored by ST and annotated with the numbers. Countries with any serotype 2 genomes are labelled.

### Ancestry analysis of GPSC96 strains of serotype 2

To elucidate the ancestry and global distribution of GPSC96 serotype 2, we performed a comprehensive phylodynamic analysis using BEAST. The resulting maximum clade credibility tree was annotated with the information on country of isolation, GPSC, ST and predicted AMR susceptibility patterns (Fig 6). The time to most recent common ancestor (tMRCA) was estimated to be 1893 AD, with a 95% highest posterior density (HPD) interval 1859-1923, further supporting the circulation of GPSC96 in the early 20^th^ century. Within this context, distinct MLST lineages showed evidence of sequential emergence: ST3744 diverged from other lineages in late 19^th^century, followed by the splits of ST74 and ST1504 in the early 20th century. More recently, newer lineages such as ST9476, ST5199, and ST5083 have emerged within ST74 since the 1960’s, indicating ongoing diversification of serotype 2 within the GPSC96 background. Collectively, these results highlight both the deep-rooted ancestry of GPSC96 and the continuous emergence of genetically distinct STs across various global contexts.

**Figure 6:**
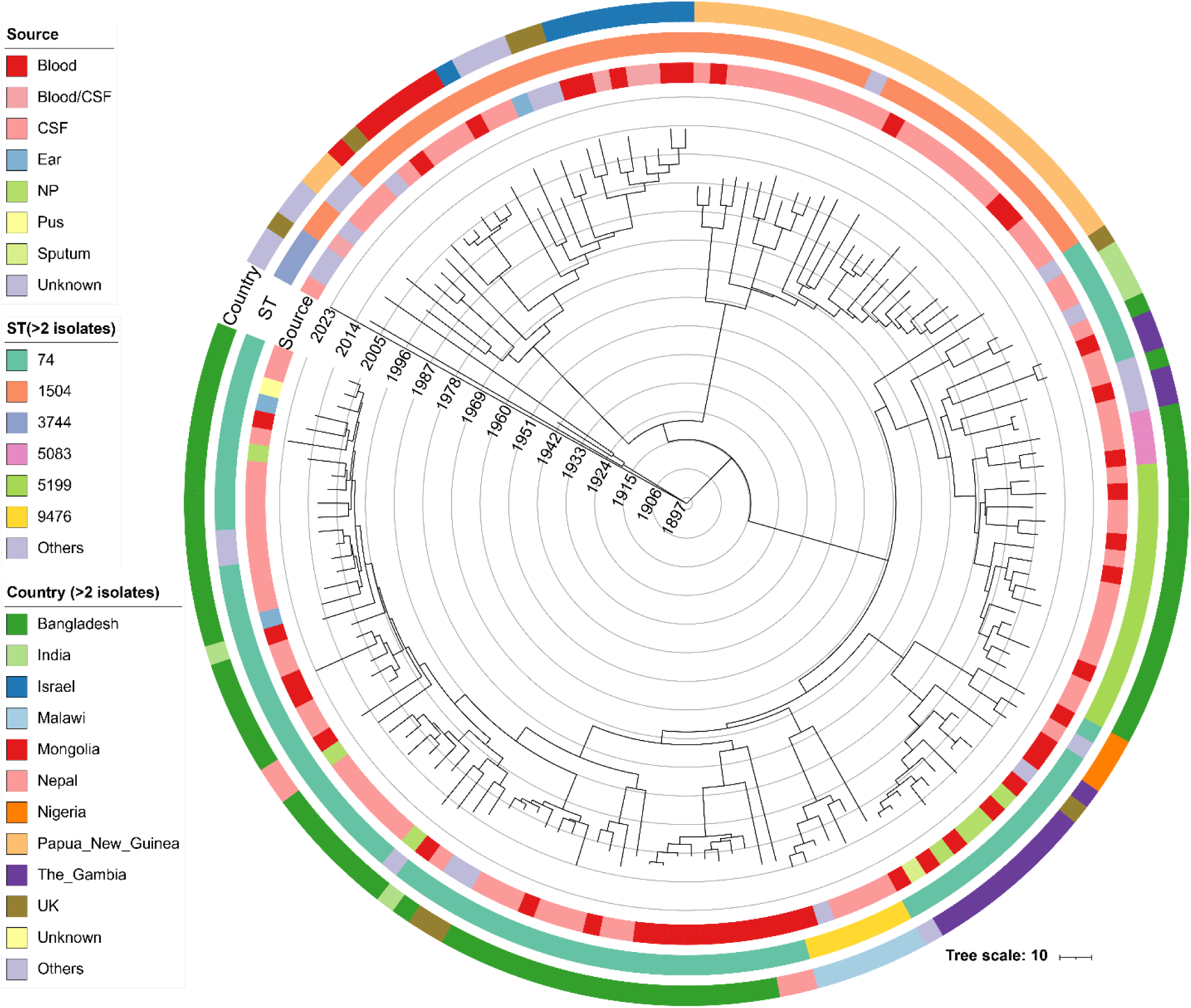
Phylodynamic analysis of global serotype 2 GPSC96 isolates. The source of isolation, ST and country of origin are shown in circles.

## Discussion

*S. pneumoniae* serotype 2 is a major contributor to non-vaccine invasive bacterial diseases in Bangladesh, primarily impacting younger children with a median age of three months. In contrast to findings from Israel^41^, our results suggest that serotype 2 isolates in Bangladesh are not linked to a single outbreak; instead, serotype 2 is circulating locally and remains an endemic cause of IPD infections in Bangladesh. This serotype is primarily linked to meningitis. Research conducted in Bangladesh has demonstrated that pneumococcal meningitis is also associated with increased rates of long-term neurodevelopmental sequelae^45^, suggesting that the socio-economic burden of serotype 2 is likely significant. Further studies could research the sequelae due to serotype 2 to quantify the true socio-economic burden of this serotype.

In our dataset, serotype 2 was found to have a high invasive potential. We compared the frequency of various serotypes in IPD relative to their prevalence in OM and nasopharyngeal carriage, finding that serotype 2 ranks among the highest invasive potential relative to other serotypes, including several already included in existing conjugate vaccine formulations. Previous studies have compared isogenic strains of *S. pneumoniae* with serotype 2 capsules to those with capsules from other serotypes (4, 6A, 7F, 14 and 19F) using murine models of infections. These studies indicated that serotype 2 capsules demonstrated lower colonization and transmission rates than the other tested serotypes^46^. Most animal studies on infections caused by serotype 2 have utilized strain D39, which belongs to GPSC622 and was isolated in 1917, subsequently grown in laboratories across the globe^47^. The higher invasive potential of serotype 2 strains observed in our study may be due to other genetic factors present in the currently circulating GPSC96 strains. Further studies can explore the molecular basis for the observed neuroinvasive nature of GPSC96 serotype 2 strains.

The detection of serotype 2 is likely underestimated for several reasons. First, diagnostic challenges may arise due to high rates of antibiotic use prior to hospital admission in countries such as Bangladesh, combined with the general antimicrobial susceptibility of serotype 2 isolates, as predicted through whole-genome sequencing. This may lead to a predominance of samples yielding negative results, with fewer positive cultures obtained. Second, in our surveillance in Bangladesh *S. pneumoniae* detection largely relied on ICT, which is approved for CSF specimens but not applicable to blood samples and is not widely used in many countries^23^. Third, some countries conduct surveillance through nasopharyngeal sampling to identify circulating serotypes^48,49^. However, our study showed that serotype 2 is rarely detected in the nasopharynx or in cases of OM. Overall, our findings imply that the actual global burden of serotype 2 is likely higher than what is currently documented in the literature.

Genomic analysis demonstrated that all serotype 2 strains sequenced from across the globe, including those from Bangladesh, have belonged to GPSC96 since the 1980s. The three genomes identified prior to the 1980s belonged to three distinct GPSCs: 96, 622 and 49, with GPSCs 49 and 622 likely being extinct. BEAST analysis suggested that GPSC96 likely emerged in the late 19^th^ or early 20^th^ century. The genomic diversity amongst serotype 2 isolates across diverse geographical regions is far less than other pneumococcal serotypes such as serotype 1 or 19A^42^.

Our study has several limitations. First, the epidemiological data were exclusively collected from Bangladesh Shishu Hospital and Institute, and there was no active surveillance system in place prior to 2004. Furthermore, genomic data were only available for 68 out of 148 samples, primarily because the remaining samples were detected using culture-negative methods. This limitation affects the completeness of our genomic analysis and restricts the strength of our conclusions. Second, we did not collect clinical features amongst meningitis cases, such as convulsions, intense irritability, lethargy etc. to know if these differ between serotype 2 vs non-serotype 2 cases. Third, the genomic data in this study was a convenience sample restricted to GPS and PubMLST database. The lack of surveillance for IPD in many countries could result in missing some genetic diversity amongst serotype 2 strains. Fourth, our data on OM and nasopharyngeal carriage were only available for the periods 2014-2019 and 2014-2016, respectively, which means our comparisons of the invasive potential for serotype 2 isolates are confined to these timeframes. However, given the substantial number of isolates from ear swabs and nasopharyngeal samples, we believe our findings are likely to be reliable.

Despite these limitations, the data collectively highlight the importance of ongoing surveillance for serotype 2 and consideration for its inclusion into newer vaccine formulations. There has been promising progress in vaccine development, with new pneumococcal conjugate vaccine formulations - PCV24 (Pn-MAPS24v/AFX3772, GSK, UK), PCV24 (VAX-24, Vaxcyte, USA), PCV25 (IVT PCV-25, Inventprise, USA) and PCV31 (VAX-31, Vaxctye, USA) - that include serotype 2 currently undergoing clinical trials^14,50^. These vaccines could represent an important advancement in controlling infections caused by serotype 2, particularly among young children who are most vulnerable to this highly invasive pneumococcal lineage.

## Acknowledgement

We thank the study participants and their parents or legal guardians, as well as the physicians and nursing staff involved in patient care and surveillance activities. We are grateful to Ms. Afroza Akter Tanni, Ms. Sharmistha Goswami, Ms. Tasnim Jabin, Mr. Mobarok Hossain, Mr. Preonath Chandrow Dev, Mr. Neoyman Nasir Shorkar and Mr. Dipu Chandra Das (Child Health Research Foundation, CHRF) for their contributions to pneumococcal culture and whole-genome sequencing.

The findings and conclusions in this manuscript are those of the authors and do not necessarily represent the official position of the U.S. Centers for Disease Control and Prevention.

## Data availability

The raw reads (both Illumina and ONT) of pneumococcal serotype 2 isolates from CHRF-PARSE and GPS2 projects supporting the conclusions of this article are available in the European Nucleotide Archive (ENA) under study accession ERP165169 (www.ebi.ac.uk/ena/browser/view/PRJEB81323). Raw reads of other isolates from GPS project are available in ENA under study accession ERP001505 (www.ebi.ac.uk/ena/browser/view/PRJEB3084). Genome assemblies and metadata are available in Supplementary Data 1. Public genomes were retrieved from Monocle and PubMLST as described in Methods.

## Ethical approval statement

Ethical approval for surveillance activities was obtained from the Ethical Review Committee of Bangladesh Shishu Hospital and Institute. Written informed consent was obtained from parents or legal guardians of all participants.

## Author Contributions Section

S.K.S. and S.S. conceived and designed the study; Y.H., A.M.T., K.B.P. and N.K. performed data analysis; B.H., S.Sa. and M.J.U. implemented surveillance and sample collection; H.R., H.N., R.M. and M.H. did laboratory testing and serotyping; R.M., D.P.K. and S.D.N. performed whole-genome sequencing; Y.H., A.M.T., H.C.H.H. and S.W.L. performed computational and phylogenomic analyses; K.P.K., M.S., L.M., S.D.B., S.S. and S.K.S. supervised the study; Y.H., A.M.T., S.W.L., S.S. and S.K.S. wrote the original draft of the manuscript; All authors reviewed and approved the final manuscript.

## Ethics declarations

### Ethics approval and consent to participate

All study protocols received approval from the Ethical Review Committee (ERC) of Bangladesh institute of Child Health (BICH) at Dhaka, Bangladesh (BICH-ERC-2/5/2011 on 05 October 2011, BICH-ERC-1/1/2025, BICH -ERC-3/1/2015 on 19 March 2015, and BICH-ERC-02/02/2021 on 03 February 2021).

Informed written consent and clinical information were obtained from parents/legal guardians for all enrolled participants in the invasive disease, and otitis media studies. Nasopharyngeal swab samples were collected with written informed consent, and only non-identifiable routine clinical data were retrospectively included.

