## Supplementary Information for "Reemergence and global distribution of an invasive lineage of *Streptococcus pneumoniae* serotype 2"

### **for**

### **Hooda et al**

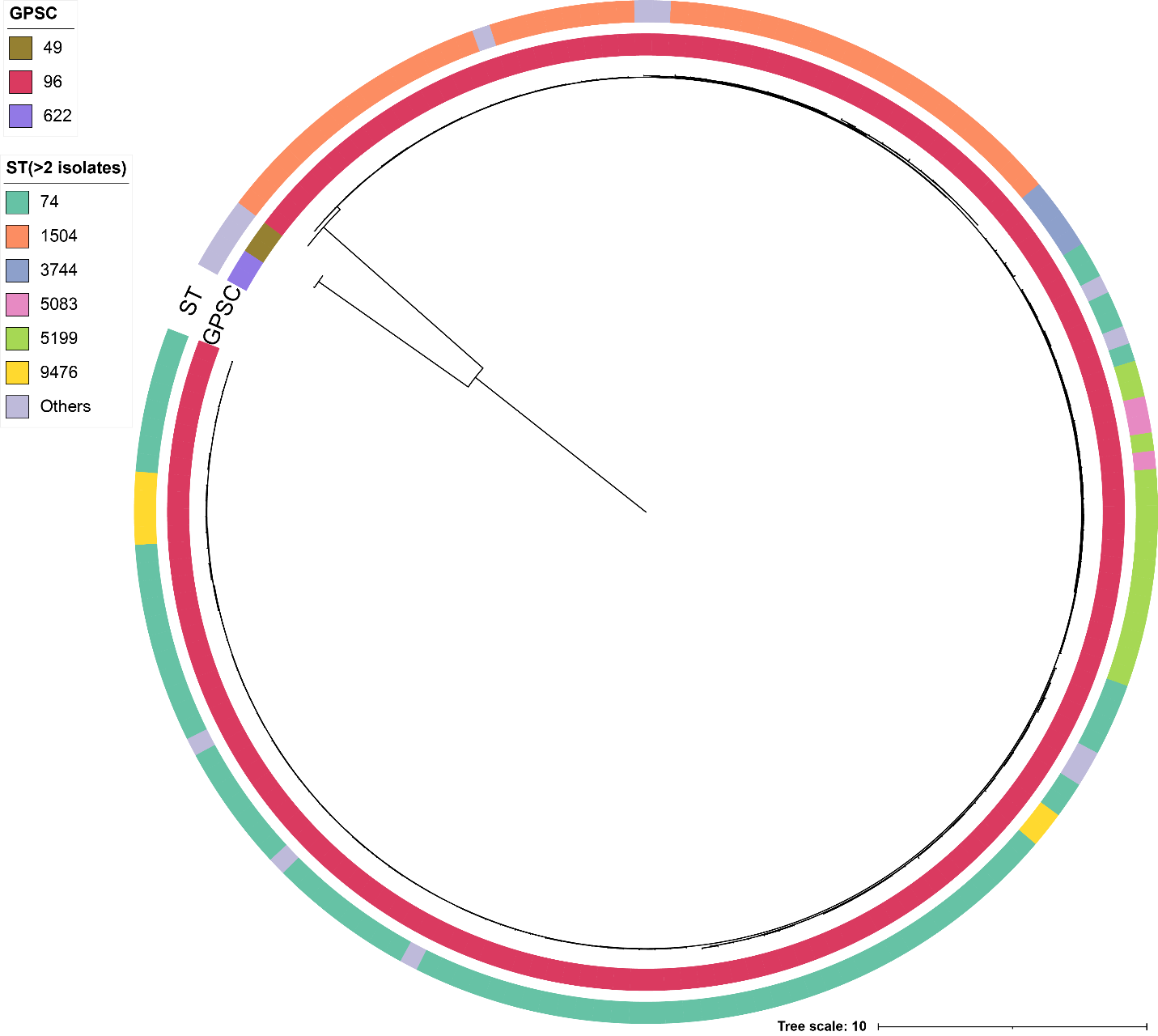

**Supplementary Figure 1.** Global pneumococcal sequence clusters (GPSCs) identified among serotype 2 isolates. Three distinct GPSCs were observed: GPSC49 (n = 2, ST574), GPSC96 (n = 166), and GPSC622 (n = 2, ST595), with GPSC96 representing the predominant cluster.

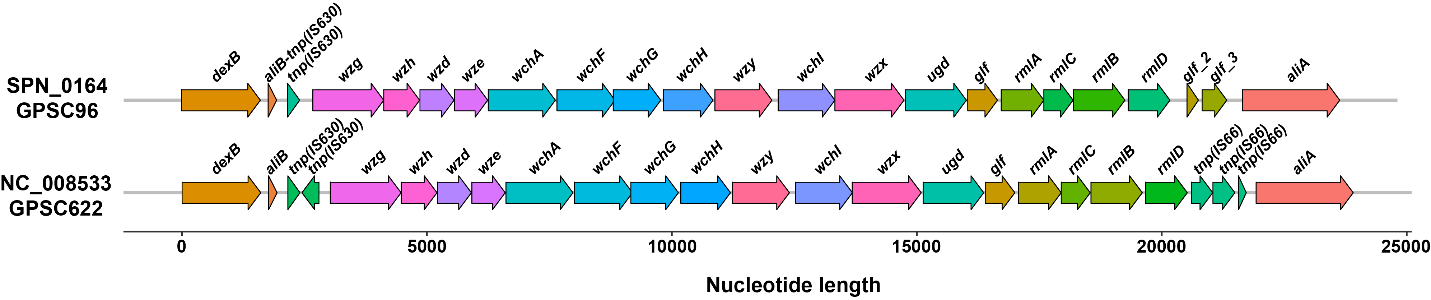

**Supplementary Figure 2. Comparison of *cps* loci between GPSC96 and GPSC622**. It shows conserved gene content but structural differences in transposon regions adjacent to *dexB* and *aliA*. The GPSC96 locus lacks a *tnp630* transposon next to *dexB*, and three *IS66* elements near *aliA* in GPSC622 are replaced by pseudogenes *glf_2* and *glf_3* in GPSC96.

**Supplementary Data 1: Metadata file of all serotype 2 genomes used in the manuscript.**
